# Quantifying the impact of quarantine duration on COVID-19 transmission

**DOI:** 10.1101/2020.09.24.20201061

**Authors:** Peter Ashcroft, Sonja Lehtinen, Daniel C. Angst, Nicola Low, Sebastian Bonhoeffer

## Abstract

The numbers of confirmed cases of severe acute respiratory syndrome coronavirus 2 (SARS-CoV-2) infection are increasing in many places. Consequently, the number of individuals placed into quarantine is increasing too. The large number of individuals in quarantine has high societal and economical costs. There is ongoing debate about the duration of quarantine, particularly since the fraction of individuals in quarantine who eventually test positive is perceived as being low. We present a mathematical model that uses empirically determined distributions of incubation period, infectivity, and generation time to quantify how the duration of quarantine affects transmission. We use this model to examine two quarantine scenarios: traced contacts of confirmed SARS-CoV-2 cases and returning travellers. We quantify the impact of shortening the quarantine duration in terms of prevented transmission and the ratio of prevented transmission to days spent in quarantine. We also consider the impact of i) test-and-release strategies; ii) reinforced hygiene measures upon release after a negative test; iii) the development of symptoms during quarantine; iv) the relationship between quarantine duration and adherence; and v) the fraction of individuals in quarantine that are infected. When considering the ratio of prevented transmission to days spent in quarantine, we find that the diminishing impact of longer quarantine on transmission prevention may support a quarantine duration below 10 days. This ratio can be increased by implementing a test-and-release strategy, and this can be even further strengthened by reinforced hygiene measures post-release. We also find that unless a test-and-release strategy is considered, the fraction of individuals in quarantine that are infected does not affect the optimal duration of quarantine under our utility metric. Ultimately, we show that there are quarantine strategies based on a test-and-release protocol that, from an epidemiological viewpoint, perform almost as well as the standard 10 day quarantine, but with a lower cost in terms of person days spent in quarantine. This applies to both travellers and contacts, but the specifics depend on the context.

## 1 Introduction

Quarantining individuals with high risk of recent infection is one of the pillars of the non-pharmaceutical interventions to control the ongoing severe acute respiratory syndrome coronavirus 2 (SARS-CoV-2) pandemic (Kucharski et al., 2020). Owing to the large fraction of transmission of SARS-CoV-2 that is pre-symptomatic or asymptomatic (Ashcroft et al., 2020; Buitrago-Garcia et al., 2020; Ferretti et al., 2020a; He et al., 2020), quarantine can prevent a substantial fraction of onward transmission that would not be detected otherwise. In mathematical modelling studies, it was estimated that thermal screening at airports would allow more than 50% of infected travellers to enter the general population (Quilty et al., 2020a; Gostic et al., 2020), which could have been prevented by mandatory quarantine. With the high or increasing case numbers that are observed in many places around the globe, however, more and more people are being placed into quarantine.

There is ongoing public debate about the appropriateness of quarantine and its duration. Quarantine lowers onward transmission in two ways: first, preventing transmission prior to symptom onset (with the assumption that symptomatic individuals will isolate) and decreasing overall transmission from persistently asymptomatic individuals. The appropriate length of quarantine thus depends on both incubation period and the temporal profile of infectiousness. In theory, quarantine periods could be avoided altogether through widespread and regular testing programmes, but the low sensitivity of reverse transcriptase PCR (RT-PCR) tests, particularly in early infection (Kucirka et al., 2020), and limitations on testing capacity in most countries, preclude this approach. Quarantine has high economic, societal and psychological costs (Nicola et al., 2020; Brooks et al., 2020). It restricts individual freedoms (Parmet & Sinha, 2020), although the level of restriction imposed is generally judged to be proportionate, given the severity of coronavirus disease 2019 (COVID-19). The low number of individuals placed in quarantine that turn out to be infected is another argument that is given against quarantine.

Individuals are generally placed into quarantine for one of two reasons: either they have been identified as a recent close contact of a confirmed SARS-CoV-2 case by contact tracing, or they have returned from recent travel to a high-risk area with levels of community transmission that are higher than in the home country (WHO, 2020). These groups of quarantined individuals differ in two important ways: compared with traced contacts, travel returners may have lower probability of being infected and have less precise information about the likely time of exposure. This raises the question whether the duration of quarantine should be the same for these two groups of individuals.

To our knowledge there are no clinical trials published that directly assess the impact of duration of quarantine on SARS-CoV-2 transmission (Nussbaumer-Streit et al., 2020). In this study, we present a mathematical model that allows quantification of the effects of changing quarantine duration. We use the distributions of incubation time (time from infection to onset of symptoms), infectivity (infectiousness as a function of days since symptom onset), and generation time (difference of time points of infection between infector and infectee). These distributions have been estimated by Ferretti et al. (2020a), combining multiple empirical studies of documented transmission pairs (Ferretti et al., 2020b; Xia et al., 2020; Cheng et al., 2020; He et al., 2020).

Using the model, we explore the effect of duration of quarantine for both traced contacts of confirmed SARS-CoV-2 cases and for returning travellers on the fraction of prevented onward transmission. We assess the effects of test-and-release strategies and the time delay between test and result. These considerations are particularly important given that multiple testing has been shown to be of little benefit (Clifford et al., 2020). We also address the role of pre-symptomatic patients becoming symptomatic and therefore being isolated independent of quarantine. Furthermore, as one of the arguments for shortening the duration of quarantine is to increase the number of people complying with the recommendation, we investigate by how much adherence needs to increase to offset the increased transmission due to earlier release from quarantine. Finally, we assess the role of reinforced individual-level prevention measures, such as mask wearing, for those released early from quarantine.

Making policy decisions about the duration of quarantine fundamentally requires specifying how the effectiveness of quarantine relates to its costs. The effectiveness can be measured in terms of the overall reduction of transmission, while economical, societal, and individual costs are likely a function the days spent in quarantine. In addition to the epidemiological outcome, which considers only the reduction in transmission, we also present results based on the ratio of transmission prevented to the average number of days spent in quarantine.

## 2 Methods

### 2.1 Quantifying the benefit of quarantine

Our primary goal is to quantify how much transmission is prevented by quarantining individuals who have been potentially exposed to SARS-CoV-2. To achieve this we need to know the time at which the individual was exposed (*t*_*E*_), as well as when they enter (*t*_*Q*_) and are released from (*t*_*R*_) quarantine (Fig. 1A). The timing of onward transmission (Fig. 1B) is determined by: the generation time distribution describing the time interval between the infection of an infector and infectee (Fig. 2A); the infectivity profile describing the time interval between the onset of symptoms in the infector and infection of the infectee (Fig. 2B); and the incubation period distribution describing the time between the infection of an individual and the onset of their symptoms (Fig. 2C). A detailed discussion about the relationships between these distributions can be found in Lehtinen et al. (2020).

**Fig. 1.**
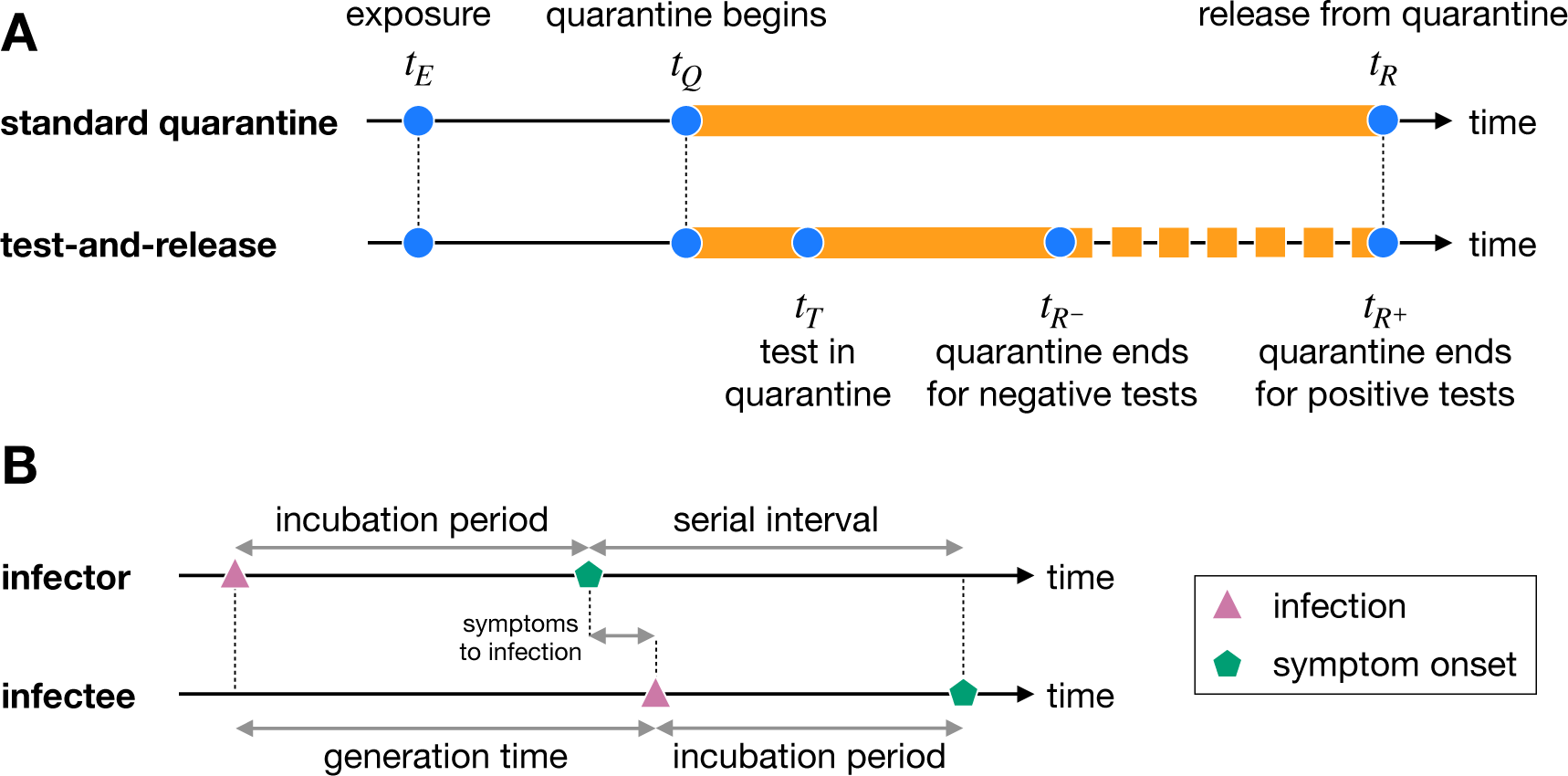
A) The timeline of quarantine. Individuals are exposed to an infector at time *t*_*E*_, and then quarantined at time *t*_*Q*_. Under the standard quarantine protocol, this individual is quarantined until time *t*_*R*_, and no onward transmission is assumed to occur during this time. Under the test-and-release protocol, quarantined individuals are tested at time *t*_*T*_ and released at time 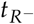 if they receive a negative test result. Otherwise the individual is isolated until 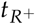. B) The timeline of infection. Here we illustrate the relationships between incubation periods, generation time, serial interval, and infection relative to symptom onset for an infector–infectee transmission pair.

**Fig. 2.**
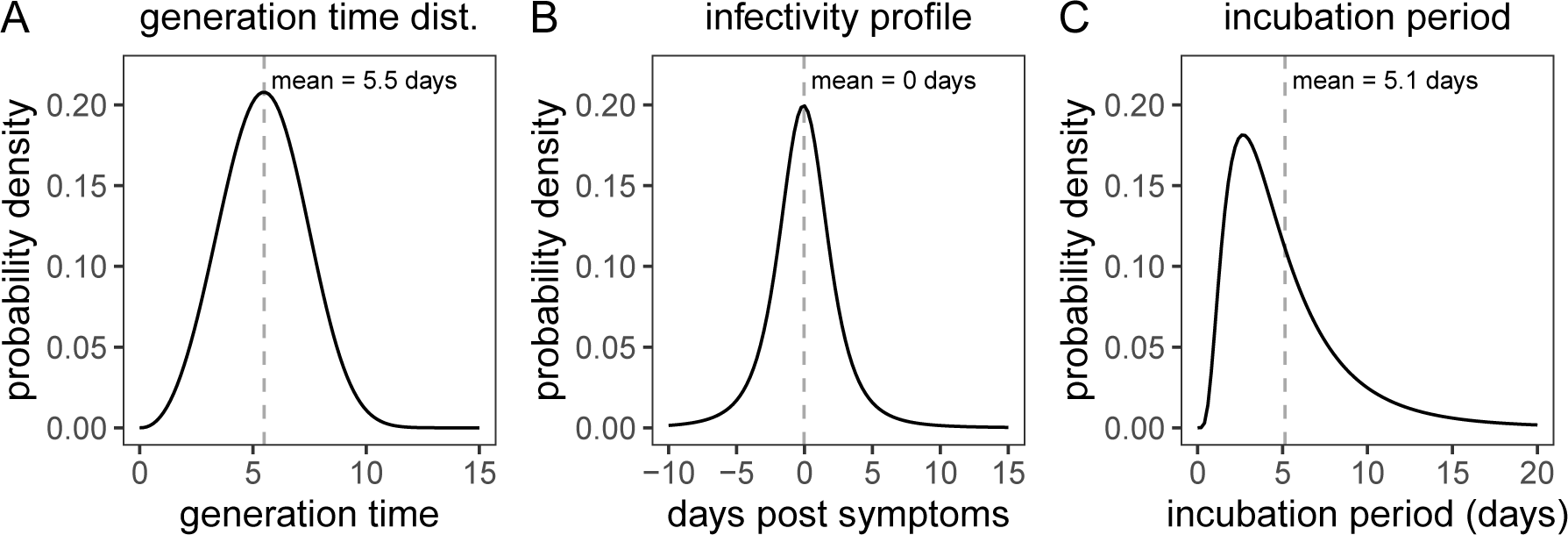
A) The generation time distribution [*q*(*t*)] follows a Weibull distribution (Ferretti et al., 2020a). B) The infectivity profile follows a shifted Student’s *t*-distribution (Ferretti et al., 2020a). C) The distribution of incubation times follows a log-normal distribution (Li et al., 2020).

Ultimately, the fraction of transmission prevented by the quarantine of an infected individual is the area under the generation time distribution *q*(*t*) [Fig. 2A] (or alternatively under the infectivity profile, Fig. 2B) between the times at which the individual enters and leaves quarantine (Grantz et al., 2020). Here we use the generation time distribution, such that the fraction of transmission prevented is

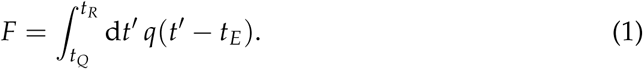

### 2.2 Traced contacts versus returning travellers

We consider the scenarios of a traced contact and a returning traveller differently, because the values of *t*_*E*_, *t*_*Q*_, and *t*_*R*_ are implemented differently in each case.

#### Traced contacts

Following a positive test result, a confirmed index case has their recent close contacts traced. From contact tracing interviews, we know when these traced contacts were last exposed to the index case (*t*_*E*_) relative to the symptom onset of the index case (*t* = 0). The contacts are then placed into quarantine at time *t*_*Q*_ = Δ_*Q*_, where Δ_*Q*_ is the sum of the delay to the index case receiving a positive test result after developing symptoms and the further delay to tracing the contacts. Under the standard quarantine procedure, the traced contacts are quarantined until day *t*_*R*_ = *t*_*E*_ + *n*, i.e. they are quarantined until *n* days after their last exposure. Note that the time spent in quarantine is *t*_*E*_ + *n* − Δ_*Q*_, which is shorter than *n*.

#### Returning travellers

Unlike our traced contacts, we do not know when travellers were (potentially) exposed. This means that quarantine starts from the date that they return (*t*_*Q*_ = 0) and lasts for *n* days until time *t*_*R*_ = *n*. For simplicity, we assume a traveller was infected at some time over a multi-day travel period −*y ≤ t*_*E*_ *≤* 0, where *y* is the duration of travel. For each exposure time *t*_*E*_, we compute the fraction of *local* transmission that would be prevented by quarantine and then compute the average over the travel duration:

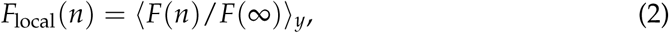

where *F*(*n*) is given by Eq. (1) and ⟨·⟩_*y*_ represents the average over the different exposure times −*y ≤ t*_*E*_ *≤* 0.

### 2.3 Fraction of individuals in quarantine that are infected

Quarantine is applied pre-emptively, such that we do not know the infection status of individuals when they enter quarantine. If only a fraction *s* of the individuals placed in quarantine are infected, then the average reduction in transmission across all individuals in quarantine is *sF*.

### 2.4 Test-and-release

The test-and-release strategy uses virological testing during quarantine to release individuals with a negative test result and to place those with a positive test result into isolation. As illustrated in Fig. 1A, a test is issued at time *t*_*T*_ *≥ t*_*Q*_. If the test is negative, the individual is released when the test result arrives at time 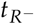. Otherwise, the individual remains in isolation until time 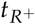. One challenge with this strategy is the high probability of a false-negative RT-PCR test result (i.e. an infected individual is prematurely released into the community). As reported by Kucirka et al. (2020), the false-negative rate is 100% on days 0 and 1 post infection, falling to 67% (day 4), 38% (day 5), 25% (day 6), 21% (day 7), 20% (day 8), and 21% (day 9), before rising to 66% on day 21. We label this function *f* (*x*), the false-negative probability on day *x* after infection. The fraction of transmission prevented by quarantine under the test-and-release strategy is:

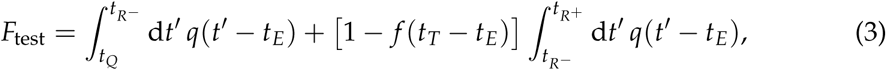

where the first term captures that all individuals are quarantined until at least the test result day 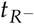, and the second term accounts for transmission prevented by remaining in quarantine until 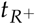 following a positive test. Again Eq. (3) is conditional on the quarantined individual being infected, and the average effect across all quarantined individuals is *sF*_test_, where *s* is the fraction of individuals in quarantine that are infected.

### 2.5 Reinforced prevention measures after early release

We further consider the possibility that individuals released after a negative test are asked to strengthen strict hygiene, mask wearing, and social distancing protocols until 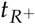. We assume that these practices reduce transmission by a fraction *r*, such that the onward transmission prevented by quarantining an infected individual is

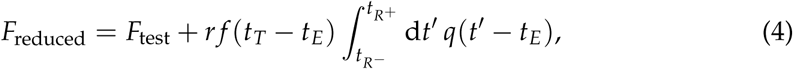

where the extra term is the transmission prevented by the reinforced prevention measures when an infected individual is prematurely released from quarantine.

### 2.6 Transmission reduction versus days spent in quarantine

One possible metric to relate the effectiveness of quarantine to its negative impact on society is to consider the ratio between the amount of overall transmission prevented and the number of person days spent in quarantine. We refer to this ratio as the utility of quarantine. Concretely, for an efficacy *F*, fraction of individuals in quarantine that are infected *s*, and average time spent in quarantine *T*, we define the utility as

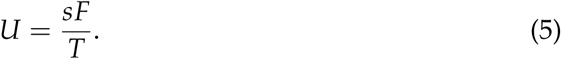

### 2.7 Adherence to quarantine

For a quarantine duration of *n* days, a fraction *α*(*n*) of individuals will adhere to the strategy, while a fraction 1 − *α*(*n*) will ignore the guideline. The fraction of transmission prevented due to quarantine is *sα*(*n*)*F*(*n*). Again *s* is the fraction of individuals in quarantine that are infected, which we assume is independent of *n*. We expect *α*(*n*) to be a decreasing function of *n*, i.e. longer quarantines have a lower adherence. For two quarantine strategies with durations *n* and *n′* to have the same overall efficacy, the adherences must satisfy *sα*(*n*)*F*(*n*) = *sα*(*n′*)*F*(*n′*), or

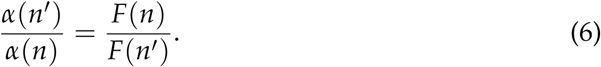

In other words, the change in the fraction of transmission prevented by quarantine must be compensated by an inverse change in the adherence. Unless otherwise stated, we assume 100% adherence.

### 2.8 Persistently asymptomatic infections

If an individual develops symptoms and ultimately tests positive while in quarantine, we can remove them from the population as they would have to isolate themselves. Importantly, this individual would be removed from the population regardless of quarantine, so the reduction of cases due to their isolation should not be counted towards the efficacy of quarantine.

Let *a* be the fraction of asymptomatic cases, who will be quarantined using the standard strategy from time *t*_*Q*_ until *t*_*R*_. We assume that the symptomatic cases would anyway be isolated once they develop symptoms (at time *t*_*S*_ as described by the incubation period distribution, Fig. 2C), so these individuals are effectively quarantined until min(*t*_*R*_, *t*_*S*_). We further assume equal transmissibility of persistently asymptomatic and symptomatic infections and that both are described by the same generation time distribution. This assumption might be an overestimate as onward transmission from persistently asymptomatic cases is less than onward transmission from symptomatic cases (Buitrago-Garcia et al., 2020). For each traced contact who is put into quarantine, the fraction of infections that would be prevented by quarantine is

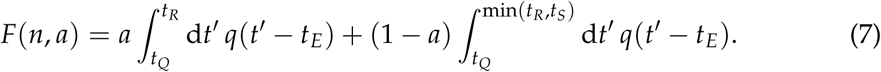

Unless otherwise stated, we assume *a* = 1.

### 2.9 Interactive app

To complement the results in this manuscript, and to allow readers to investigate different quarantine scenarios, we have developed an online interactive application. This can be found on the *CH Covid-19 Dashboard* at https://ibz-shiny.ethz.ch/covidDashboard/.

## 3 Results

### 3.1 Quarantining traced contacts of confirmed SARS-CoV-2 cases

Any shortening of a traced contact’s quarantine duration will lead to an increase in transmission from that individual, but the degree of increase depends on the extent of the shortening. The expected onward transmission (from an infected contact) that is prevented by quarantine [*F*; Eq. (1)] shows the diminishing return of increasing the quarantine duration (Fig. 3A). Furthermore, as the time delay to quarantine (Δ_*Q*_) increases, the maximum efficacy of quarantine is reduced (because infected contacts have already transmitted before being quarantined). If the duration of quarantine is longer than 10 days, then little can be gained in terms of prevention by quarantining for longer, but reducing the delay to starting quarantine does lead to increased efficacy.

Note that we have assumed that the contact was infected at the last time of exposure. If there have been multiple contacts between them and the index case, then transmission may have occurred earlier and we would overestimate the efficacy of quarantine.

**Fig. 3.**
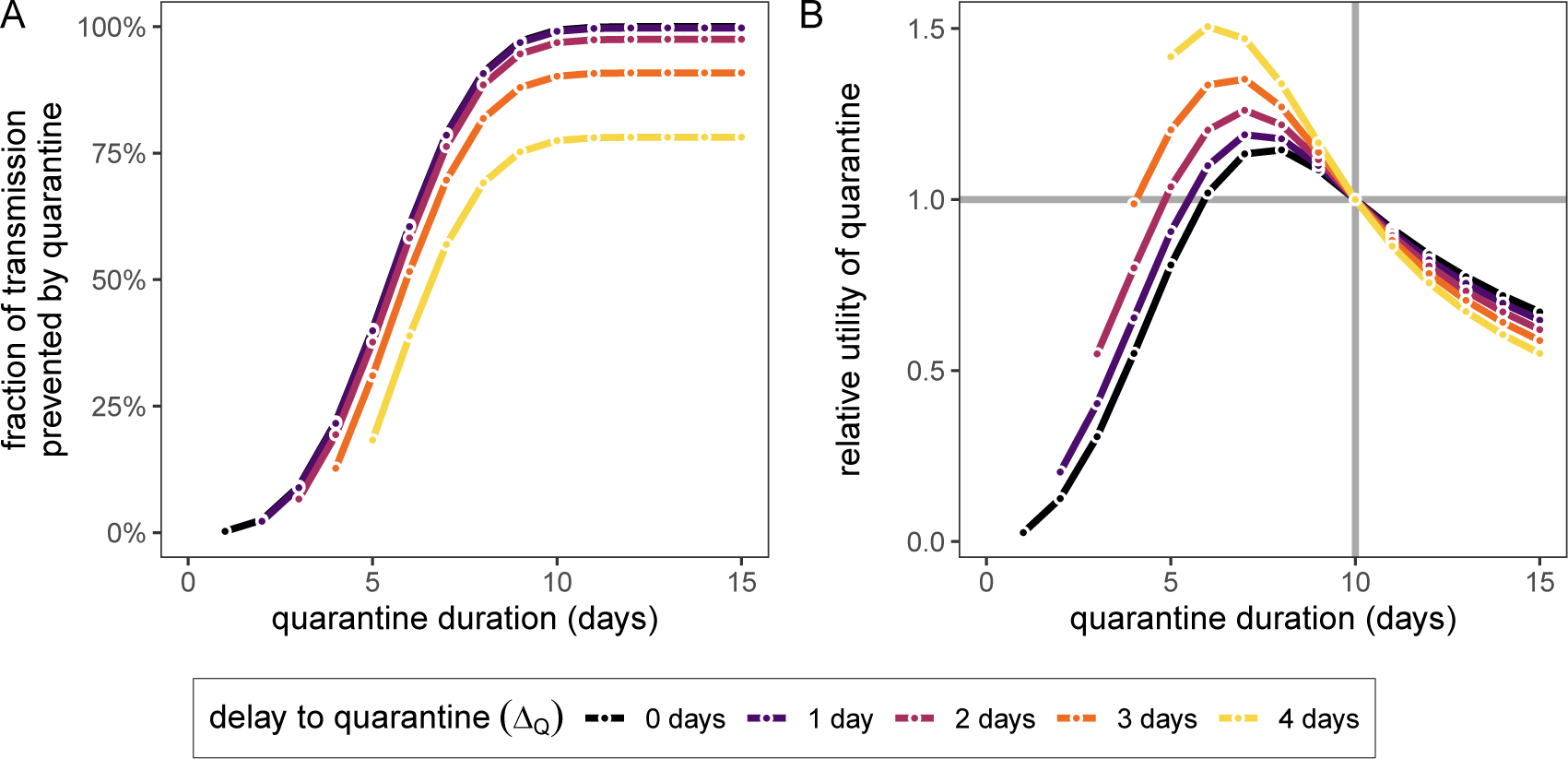
A) The fraction of total onward transmission per quarantined infected contact that is prevented by quarantine [Eq. (1)]. B) The relative utility of different quarantine durations (x-axis) compared to *n* = 10 days, i.e. *U*(*n′/*)/*U*(10) [utility as defined Eq. (8)]. Colours represent the delay to starting quarantine, Δ_*Q*_. We use *t*_*E*_ = 0, which from the infectivity profile is the mean infection time of contacts if the index case develops symptoms at *t* = 0.

The utility of the quarantine strategy, as defined in Eq. (5), is

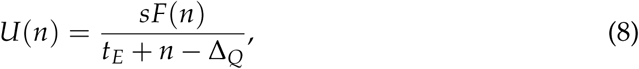

which depends on the fraction of individuals in quarantine that are infected, *s*. However, comparing two different quarantine durations *n* and *n′* through their relative utility, i.e. *U*(*n′*)/*U*(*n*), eliminates this dependence on *s*. Therefore, the argument that we should shorten quarantine because of the low probability of quarantined individuals being infected is misguided in this situation. By calculating the relative utility, we observe that there is an optimal strategy which maximises the ratio between the fraction of transmission prevented and the number of days spent in quarantine (Fig. 3B). This would be a duration of six to eight days, depending on the delay to starting quarantine Δ_*Q*_.

#### Test-and-release strategy

Quarantined individuals are tested *x* days after exposure and released if the test result is negative. As above, quarantine begins at time *t*_*Q*_ = Δ_*Q*_. The test is conducted at *t*_*T*_ = *t*_*E*_ + *x*, and the result is received after a delay Δ_*T*_ at time 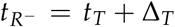. Individuals with a negative test result are released, otherwise they remain in quarantine until time 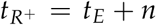. We here assume that *n* is large enough such that negligible transmission would occur after this period.

For *n* = 10, Δ_*Q*_ = 3, and *t*_*E*_ = 0, standard quarantine (without testing) would prevent 90.2% of onward transmission from an infected traced contact. This represents the maximum attainable efficacy: the onward transmission prevented under a test-and-release strategy will always be below this level (Fig. 4A). This is because of the chance of prematurely releasing an infectious individual who received a false-negative test result. On the other hand, the test-and-release strategy always performs better than not testing if the release time is the same as the quarantine duration (grey line in Fig. 4A). Hence, these scenarios provide upper and lower bounds for the efficacy of the test-and-release strategy. The efficacy of the test-and-release strategy increases if we test later in quarantine, because we not only increase the duration of quarantine but also reduce the false-negative probability.

**Fig. 4.**
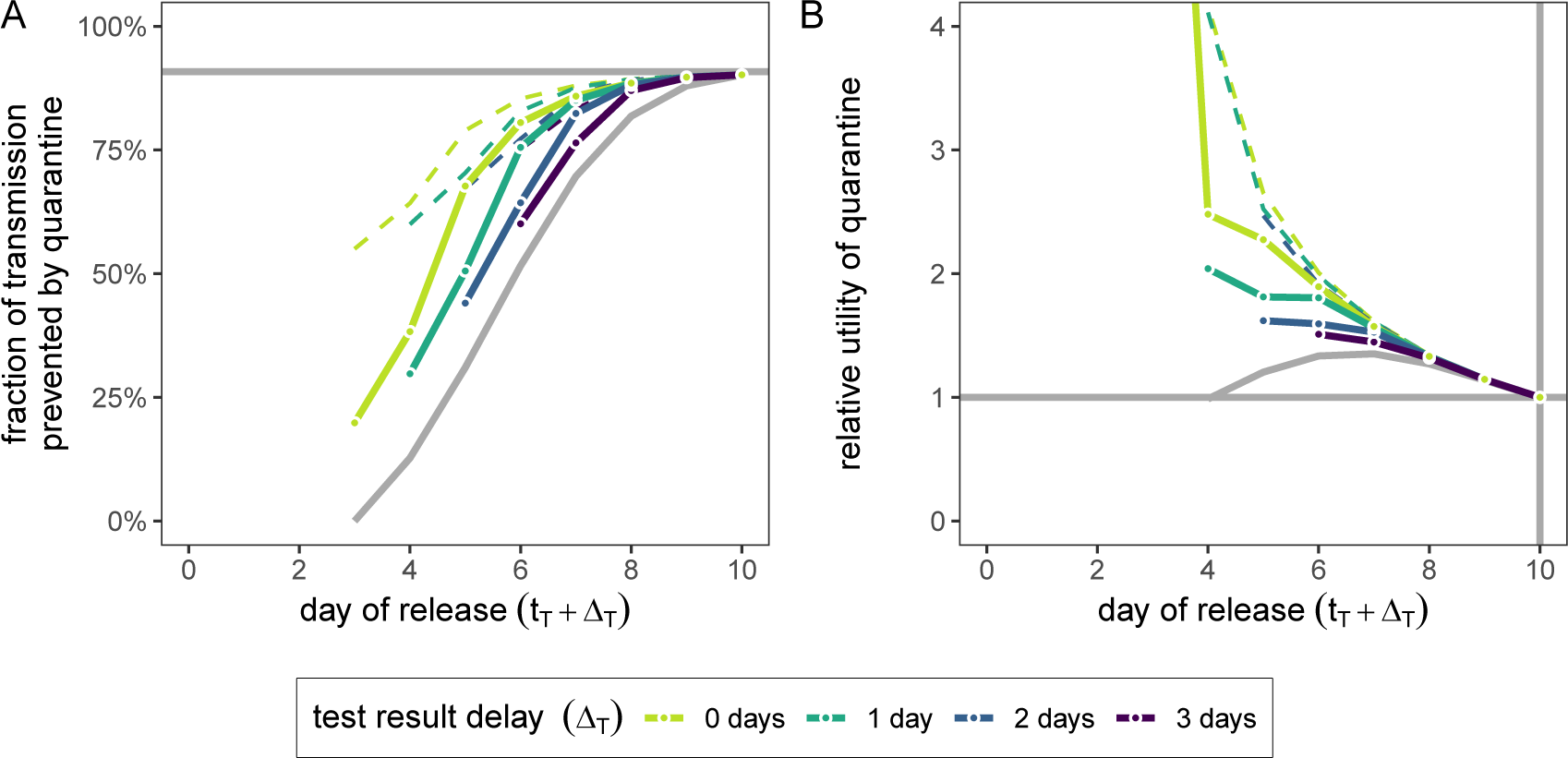
A) The impact of the test-and-release quarantine strategy, in terms of what fraction of total onward transmission per infected traced contact is prevented by quarantine [*F*_test_; Eq. (3)]. The grey lines represent the upper and lower bounds for the fraction of transmission that can be prevented by quarantine without testing [*F*; Eq. (1)]. B) The relative utility of different test-and-release quarantine durations compared to standard quarantine with duration *n* = 10 days, i.e. *U*_test_(*n′, x*)/*U*(10) [Eq. (9)]. The grey curve is the relative utility of standard quarantine without testing. In both panels we use *t*_*E*_ = 0, which from the infectivity profile is the mean infection time of contacts if the index case develops symptoms at *t* = 0, and Δ_*Q*_ = 3 as the delay until quarantine begins. Individuals are tested on day *t*_*T*_ = *x* after exposure and released on day *x* + Δ_*T*_ (x-axis) if negative (colour corresponds to Δ_*T*_). Positive testing individuals are released at day *n′* = 10. We assume that the fraction of individuals in quarantine that are infected is *s* = 0.1, and that there are no false-positive test results. Dashed lines in both panels assume the released individuals have a 50% reduced transmission (*r* = 0.5) due to reinforced hygiene and social distancing measures [Eq. (4)].

Current laboratory-based RT-PCR tests have a median turnaround of 24-48 hours. This delay is primarily operational, and so could be reduced by increasing test through-put. There are more rapid antigen-detection tests which can provide same day results, but with lower sensitivity and specificity compared to RT-PCR tests (Guglielmi, 2020). Here we assume that tests have the same sensitivity and specificity regardless of the delay to result. Comparing to a test with delay Δ_*T*_ = 2 days, we observe that using a rapid test (Δ_*T*_ = 0) can reduce the quarantine duration of individuals with a negative test result by one day while maintaining the same efficacy (Fig. 4A): the extra accuracy gained by waiting one extra day until testing balances the increased transmission caused by reducing the duration. E.g., a rapid test on day six has roughly the same efficacy (80.5%) as testing on day five and releasing on day seven (82.4%), while shortening the quarantine duration of individuals with a negative test result from seven to six days. Note that

The function *F*_test_ shown in Fig. 4A describes only the epidemiological benefit of quarantine. The average time spent in quarantine will be *t*_*E*_ + *x* + Δ_*T*_ − Δ_*Q*_ + *s*[1 − *f* (*x*)](*n* − *x* − Δ_*T*_), where only positive cases *s* have the ability to return a positive test and remain in quarantine (i.e. we assume there are no false-positive test results). Hence the utility of the test-and-release strategy, using the definition in Eq. (5), is

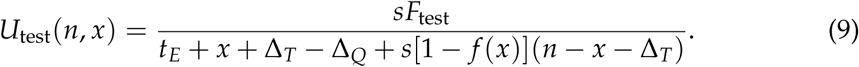

We can now compare the test-and-release strategy with standard quarantine of duration *n* = 10 days using the relative utility *U*_test_(*n′, x*)/*U*(*n*), which now depends on *s*. Based on this metric, we see that testing-and-releasing before the end of the standard quarantine period increases the utility (Fig. 4B). Testing on day five and releasing on day seven (assuming a delay Δ_*T*_ = 2 days between the test and receipt of a negative result) has a relative utility of 1.5 compared to a standard 10 day quarantine. Reducing the delay between test and result leads to a corresponding increase in utility: a rapid test (Δ_*T*_ = 0) on day six has a relative utility of 1.9 for an equivalent efficacy (Fig. 4B).

Finally, we consider the effects of reinforced prevention measures, where individuals who receive a negative test result are released from quarantine but must adhere to strict hygiene and social distancing protocols until the end of the full quarantine. Considering a 50% reduction of post-quarantine transmission, we see large increases in both efficacy and utility for early testing strategies, but with diminishing returns as the time at which tests are conducted is increased (dashed lines in Fig. 4). Note that we assume no contribution to the number of days spent in quarantine in the utility function due to mask wearing and social distancing in the post-release phase.

#### Adherence to quarantine and proportion of individuals with persistently asymptomatic infection

Shortening the duration of quarantine from 10 days to five days would require more than twice as many individuals to adhere to the quarantine guidelines in order to maintain the same overall efficacy (Fig. 5A). If quarantine is shortened further, then the required increase in adherence grows rapidly and soon becomes unfeasible (the maximum possible adherence is *α* = 1, so depending on the baseline level of adherence it may be impossible to increase this by the required factor). Hence the argument of shortening quarantine to increase adherence is of limited use.

**Fig. 5.**
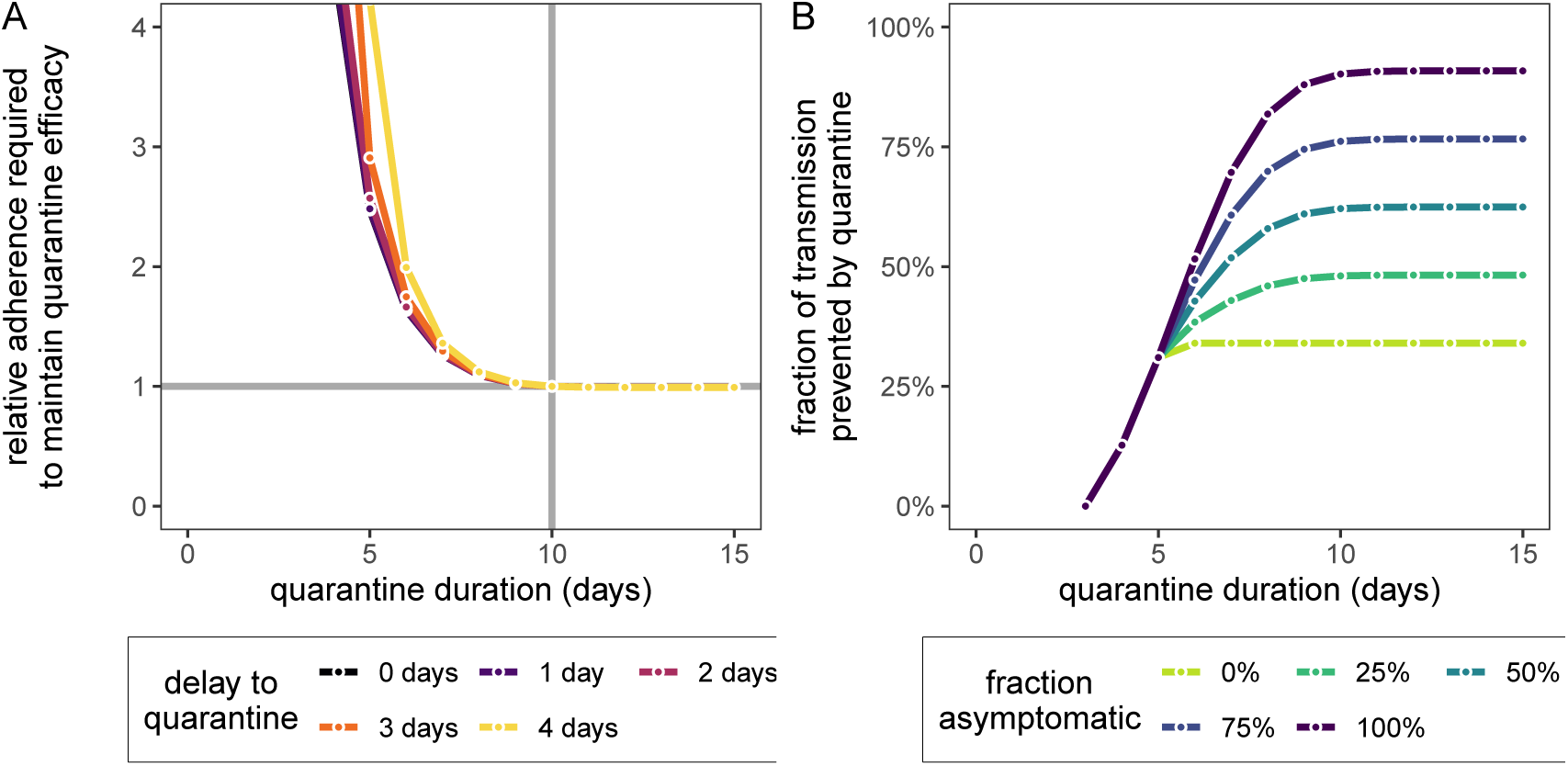
A) The change in adherence *α* needed to maintain quarantine efficacy of the *n* = 10 day strategy if we change the quarantine duration to *n′* days (x-axis), i.e. *α*(*n′*)/*α*(10) [Eq. (6)]. B) The impact of symptomatic cases on the fraction of total onward transmission per infected traced contact that is prevented by quarantine [Eq. (7)]. We fix Δ_*Q*_ = 3 as the delay until quarantine, and *t*_*S*_ = 5, which is the mean incubation time. The curve for *a* = 1 corresponds to Fig. 3A. In both panels we use *t*_*E*_ = 0, which is the mean infection time of secondary cases based on the infectivity profile.

As a final consideration, we note that our quantification of the fraction of transmission prevented by quarantine [Eq. (1)] is more relevant to individuals with persistently asymptomatic SARS-CoV-2 infection than to those who develop symptoms during the quarantine phase and are subsequently tested and isolated. The fraction of transmission prevented by quarantine is an increasing function of the fraction of asymptomatic cases (Fig. 5B). This means that we likely overestimate the efficacy of quarantine as we are also counting transmission that is prevented by isolation following a positive test result.

### 3.2 Quarantine of returning travellers

Returning travellers that have been infected on a short journey will have, on average, used up less of their infectivity potential by the time they return than a traveller who was infected on a long journey. Hence, the *total* transmission that can be prevented by a long quarantine period (i.e. 10 days) upon arrival, is greater for short trips (Fig. S1). When considering the fraction of *local* transmission that can be prevented by quarantine [*F*_local_; Eq. (2)], then shorter quarantine durations have a greater impact on long than short trips (Fig. 6A). Again, this is because, on average, the traveller on a long trip would have been exposed earlier and they will be infectious for a shorter time period after arrival. A standard quarantine duration of 10 days is enough to prevent almost all local transmission for any travel duration (Fig. 6A).

**Fig. 6.**
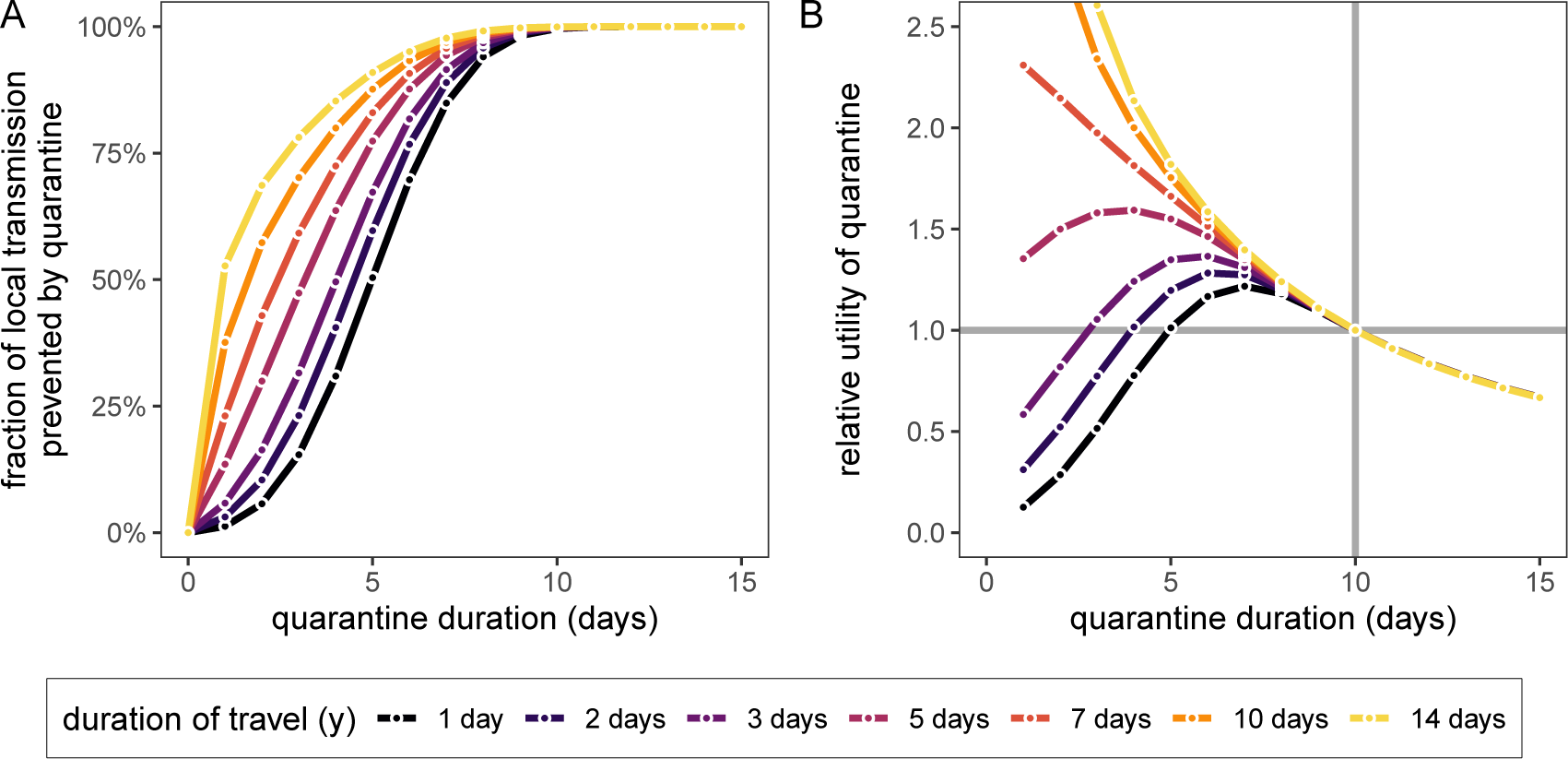
A) The fraction of local onward transmission per quarantined traveller that is prevented by quarantine [*F*_local_; Eq. (2)]. B) The relative utility of different quarantine strategies (x-axis) compared to *n* = 10 days, i.e. *U*(*n′*)/*U*(10) [Eq. (10)]. Colours represent the duration of travel *y* and we assume infection can occur on any day −*y ≤ t*_*E*_ *≤* 0 with uniform probability.

The human cost of quarantine here is *n* days, and hence the utility is simply *U*(*n*) = *sF*_local_(*n*)/*n*. Comparing two different strategies *n* and *n′*, we have the relative utility

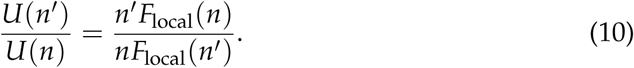

As above, an individual who has been infected during a long trip will have, on average, been infected earlier and therefore will have, on average, less remaining infectivity potential upon return compared with an individual who travelled for a short duration. Hence, if an individual traveller is to be quarantined, then the optimum duration of quarantine, based on this metric of utility, would depend on the duration of their travel, with shorter journeys requiring longer quarantine (Fig. 6B). This might be counter-intuitive because individuals who have been on longer journeys to high risk countries have a higher probability of being infected. The absolute utility *U* of quarantining such individuals is indeed higher than for individuals returning from shorter journeys. However, here, we are not considering the question of whether to quarantine or not, but we are assuming that the individual is quarantined and are trying to optimise the duration of quarantine in response to the expected infection dynamics. As the relative utility [Eq. (10)] is independent of *s*, the prevalence of disease in the travel destination (which should correlate with the fraction of travellers becoming infected at that destination) does not influence the optimal quarantine duration.

#### Test-and-release strategy

If a returning traveller is tested at time *t*_*T*_ = *x* during quarantine then they are released on day 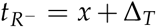 if the test is negative, or else kept in quarantine until day 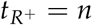. We focus on a seven day trip and fix *n* = 10 days, which is long enough to prevent almost 100% of local onward transmission from the returning traveller (Fig. 6A). The fraction of local transmission prevented by testing-and-releasing an infected traveller accounts for false-negative test results.

As was the case for the traced contacts, the fraction of local transmission prevented by standard quarantine bounds the efficacy of the test-and-release quarantine strategy (Fig. 7A). The timing of the test can have significant impact on prevented transmission; late testing reduces the false negative probability but increases the stay in quarantine. An important consequence of this is that testing on arrival (*x* = 0) is a poor strategy for limiting transmission: testing upon return at day zero only prevents 54.1% of local transmission, but testing on day five and releasing on day seven prevents 98.5% of local transmission.

**Fig. 7.**
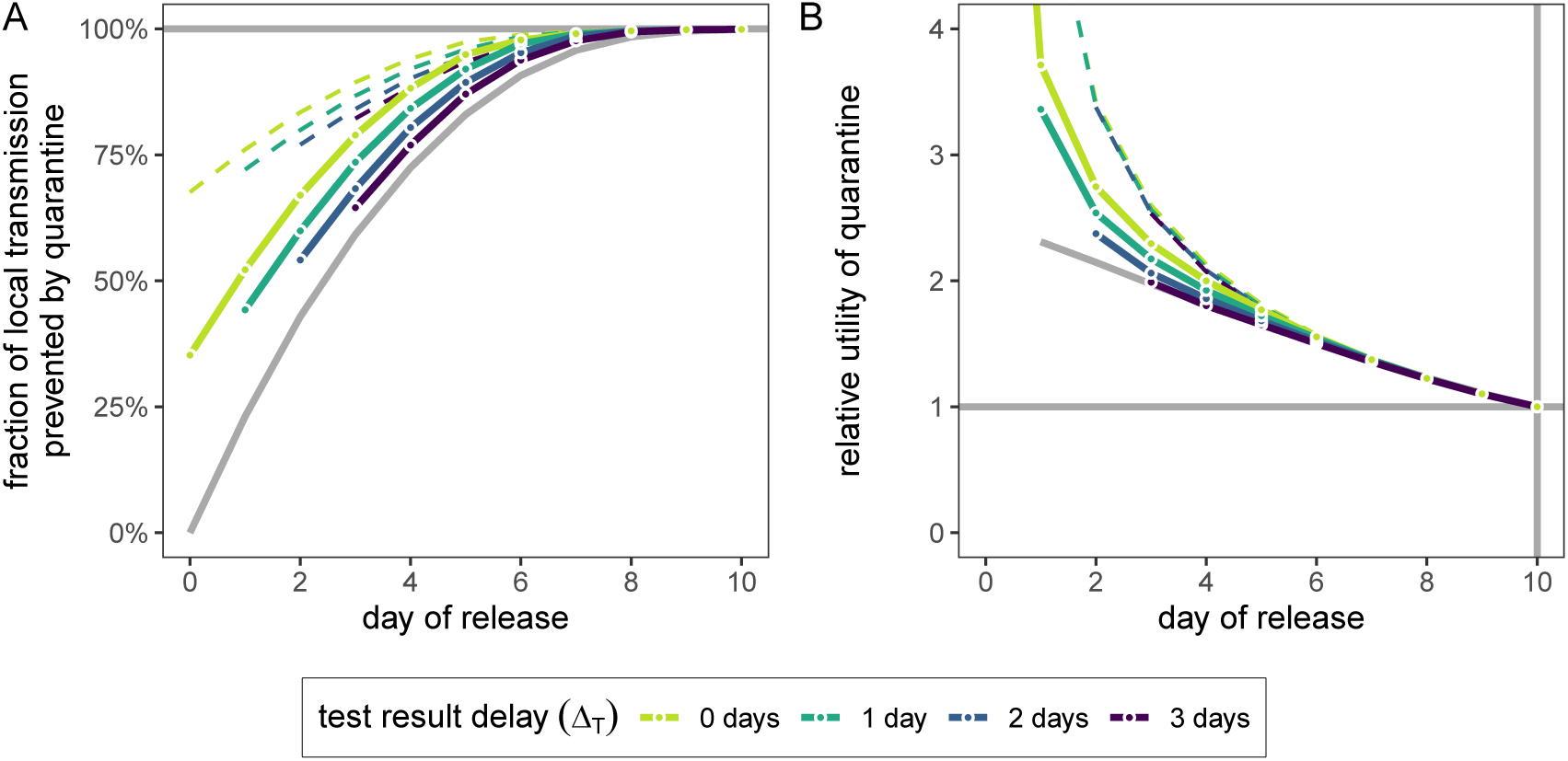
A) The impact of test-and-release for quarantined travellers, in terms of what fraction of local onward transmission per quarantined infected traveller is prevented by quarantine. The grey lines represent the upper and lower bounds for the fraction of transmission that can be prevented by quarantine without testing. B) The relative utility of different test-and-release quarantine durations compared to standard quarantine with duration *n* = 10 days. The grey curve is the relative utility of standard quarantine without testing. In both panels we consider a travel duration of *y* = 7 days and we assume infection can occur on any day −*y* ≤*t*_*E*_ ≤0 with uniform probability. Individuals are tested on day *x* after returning on day 0 and released on day *x* + Δ_*T*_ (x-axis) if negative (colour corresponds to Δ_*T*_). We assume that the fraction of individuals in quarantine that are infected is *s* = 0.1, and that there are no false-positive test results. Dashed lines in both panels assume the released travellers have a 50% reduced transmission (*r* = 0.5) due to extra hygiene and social distancing measures imposed by reduced quarantine [Eq. (4)].

A rapid test could eliminate the two-day delay between test and result. If the rapid test (Δ_*t*_ = 0) has the same sensitivity and specificity as the laboratory-based RT-PCR test, then the duration of quarantine of individuals with a negative test result can be shortened by one day with minimal loss in efficacy compared to the standard test (Fig. 7A).

Fig. 7A describes the epidemiological effect of quarantining individuals. The average duration of quarantine will be *⟨x* + Δ_*T*_ + *s*[1 − *f* (*x* − *t*_*E*_)](*n* − *x* − Δ_*T*_)*⟩*_*y*_, and hence the utility *U*_test_(*n, x*) and relative utility *U*_test_(*n′, x*)/*U*(*n*) will depend on *s*. Early testing greatly reduces the average duration of quarantine and hence leads to increased utility (Fig. 7B). Enforcing additional hygiene and social distancing guidelines following a negative test and release can increase both efficacy and utility for early testing strategies, but with diminishing returns as the release date is increased (dashed lines in Fig. 7).

We note that the relative utility of the test-and-release strategy depends on the fraction of individuals in quarantine that are infected (*s*), and this fraction may change depending on disease prevalence at the travel destination and the duration of travel. E.g., the infected fraction of travellers returning from a long stay in a high-risk country is likely to be higher than the infected fraction of travellers returning from a short stay to a low risk country. In Fig. 7B we keep *s* fixed.

Finally, we note that the change in adherence required to balance a change in efficacy for shortened quarantine durations is dependent on the travel duration, with short travel durations requiring a greater increase in adherence compared with longer travel durations (Fig. S2A).

## 4 Discussion

Quarantine is one of the most important measures in controlling the ongoing SARS-CoV-2 epidemic due to the large fraction of presymptomatic and asymptomatic transmission. A quarantine period of 10 days from exposure, as currently implemented in Switzerland, is long enough to prevent almost all onward transmission from infected contacts of confirmed cases from the point of entering quarantine: increasing the duration of quarantine beyond 10 days has no extra benefit. Reducing the delay to quarantining individuals increases the fraction of total transmission that is preventable. The same 10 day quarantine duration will prevent almost all local onward transmission from infected travel returners from the time of arrival, independent of their travel duration.

Any decrease in the duration of quarantine of an infected individual will result in increased onward transmission. Furthermore, our analyses suggest that this increase in transmission cannot realistically be compensated by increased adherence for significantly shortened quarantine (fewer than five days). However, there are diminishing returns for each day that we add to quarantine: increasing the duration from 10 days has a negligible effect in terms of reduced transmission. One therefore has to assess how much human cost, measured in terms of days spent in quarantine, we are willing to spend to prevent disease transmission. By comparing the ratio of prevented transmission to quarantine duration, we have shown that maximal utility strategies can exist. This ratio is maximised for quarantine durations of six to eight days after exposure for traced contacts, and potentially less for returning travellers depending on their duration of travel. Importantly, under this metric the fraction of individuals in quarantine that are infected does not affect the optimal duration of quarantine. Therefore, the argument that we should shorten quarantine because of the low probability of being infected is misguided under our definition of utility and in the absence of testing during quarantine.

A test-and-release strategy will lead to a lower average quarantine duration across infected and non-infected individuals. However, due to the considerable false-negative probability of the RT-PCR test (Kucirka et al., 2020), this strategy also leads to increased transmission as infectious individuals are prematurely released. Nevertheless, test-and-release strategies prevent more transmission than releasing without testing, and hence test-and-release increases the utility of quarantine. Reducing the delay between test and result leads to further reduced transmission and increased utility, and reinforcing individual prevention measures after release is also effective for short quarantine periods.

The ratio of transmission prevented versus days spent in quarantine is only one possible definition of utility. Defining the appropriate function is ultimately a policy question: the economical, societal, and individual costs are likely a function the days spent in quarantine, but we cannot determine the shape of this function. Another perspective is that the utility of preventing transmission is crucially dependent on whether it brings the effective reproductive number under one.

Ultimately, bringing the reproductive number below one requires a combination of effective measures including isolation, physical distancing, hygiene, contact tracing, and quarantine (Kucharski et al., 2020). Effective quarantine is only possible in the presence of efficient contact tracing to find the potentially exposed individuals in a short time, as well as surveillance of disease prevalence to identify high-risk travel. Further reducing the time taken to quarantine a contact after exposure and reducing the delay between test and result will allow average quarantine durations to be shorter, which increases the benefit to cost ratio of quarantine.

The scenarios of returning travellers and traced contacts of confirmed SARS-CoV-2 cases differ in the probability of having been exposed and infected and on the information available about the likely window of exposure. The impact of quarantining returning travellers depends on the duration of travel, and whether we consider the local prevention of transmission or the total transmission prevented by quarantine. However, a single test done immediately after return can only prevent a small fraction of the transmission from a returning traveller because of the false-negative rate of the RT-PCR test early in infection. Therefore testing should be postponed until as late as possible, and utilising rapid tests could be crucial if their performance characteristics are acceptable. This same principle also applies to traced contacts. Our findings are aligned with those of two recent simulation studies which estimate the role that quarantine plays in limiting transmission from returning travellers (Clifford et al., 2020) and from traced contacts (Quilty et al., 2020b).

Our results are based on the latest estimates of the generation time distribution of COVID-19 (Ferretti et al., 2020a). Potential limitations to our approach could be that these distributions may change throughout the epidemic, particularly depending on how people respond to symptoms (Ali et al., 2020). Furthermore, these distributions could be different between persistently asymptomatic and symptomatic individuals (Buitrago-Garcia et al., 2020). For travellers, another consideration is that lengthy quarantine is seen as a deterrent to travel to high risk areas (IATA, 2020). Any shortening of quarantine may lead to an increase in travel volume, potentially leading to a compounded increase in disease transmission.

In the absence of empirical data about the effectiveness of different durations of quarantine, mathematical modelling can be used objectively to explore the fraction of onward transmission by infected contacts or returning travellers that can be prevented. However, determining the optimal quarantine strategy to implement depends on the impact that shortening the duration of quarantine has on individuals, society, and the economy, versus how much weight is assigned to a consequential increase in transmission. Both the individual, societal and economic impact, as well as the weight of transmission increase, will have to be considered based on the current epidemiological situation. We have shown that there are quarantine strategies based on a test-and-release protocol that, from an epidemiological viewpoint, perform almost as well as the standard 10 day quarantine, but with a lower cost in terms of person days spent in quarantine. This applies to both travellers and contacts, but the specifics depend on the context.

## Data Availability

Code is publicly available at http://github.com/ashcroftp.

## Supplemental figures

**Fig. S1.**
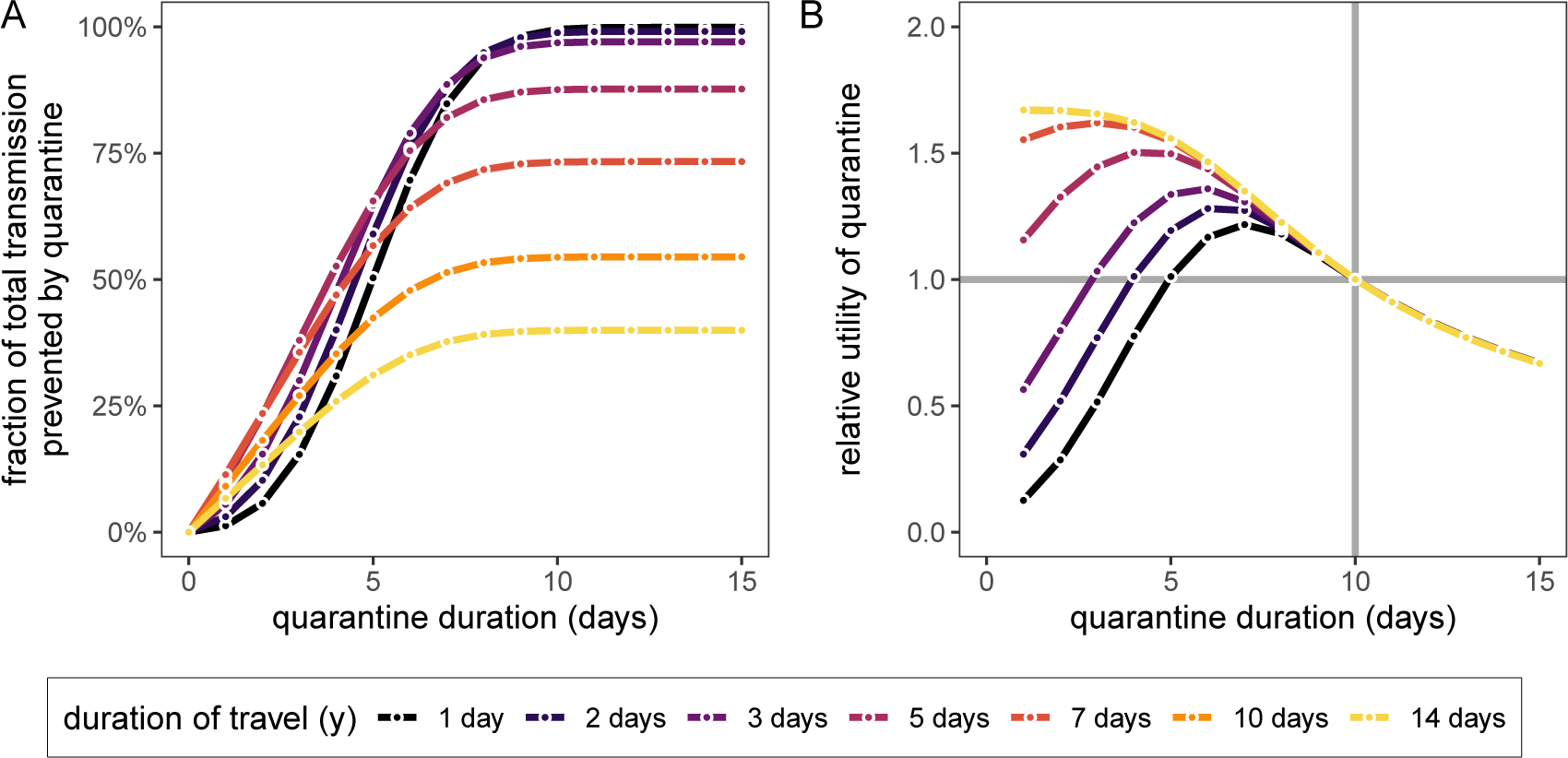
Total transmission prevented in returning travellers. A) The fraction of total onward transmission per quarantined traveller that is prevented by quarantine [*F*; Eq. (1)]. B) The relative utility of different quarantine strategies (x-axis) compared to *n* = 10 days, i.e. *U*(*n′*)/*U*(10) [Eq. (10)]. Colours represent the duration of travel *y* and we assume infection can occur on any day −*y ≤ t*_*E*_ *≤* 0 with uniform probability.

**Fig. S2.**
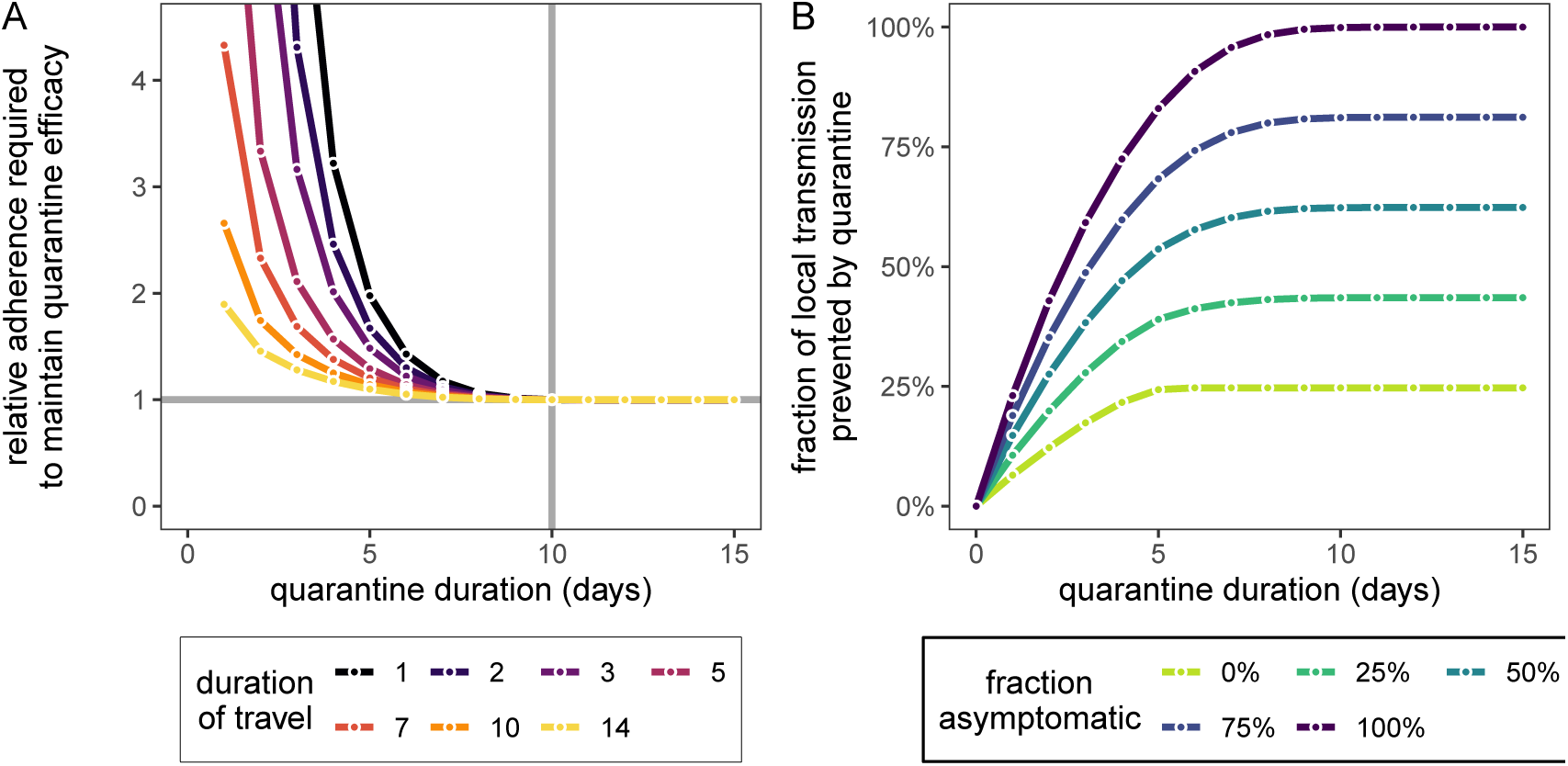
Adherence and symptoms for returning travellers. A) The change in adherence needed to maintain quarantine efficacy of the *n* = 10 day strategy if we change the quarantine duration to *n′* days (x-axis), i.e. *α*(*n′*)/*α*(10) = *F*_local_(10)/*F*_local_(*n′*). B) The impact of symptomatic cases on the fraction of total onward transmission per quarantined traveller that is prevented by quarantine. We fix the travel duration to *y* = 7 days and assume *t*_*E*_ is uniformly distributed between −*y* and 0. The curve *a* = 1 corresponds to Fig. 6. We use the mean incubation time of five days, such that *t*_*S*_ = *t*_*E*_ + 5.

## Notes

### Competing Interest Statement

The authors have declared no competing interest.

### Funding Statement

This work was supported by the Swiss National Science Foundation. The funders of the study had no role in study design, data collection, data analysis, data interpretation, or writing of the manuscript.

### Author Declarations

No IRB needed

### Summary of Updates

New authors added; Figure presentation modified; Reworked Figure 1; Figures 4 and 7 updated to include variable delay to test result; Figure 6 changed to fraction of local transmission prevented. (Original Figure 6 moved to supplement); Figure 8 moved to supplement; General improvements to text throughout.

